# SOD1-ALS-Browser: a web-utility for investigating the clinical phenotype in *SOD1* amyotrophic lateral sclerosis

**DOI:** 10.1101/2023.03.03.23286719

**Authors:** Thomas P Spargo, Sarah Opie-Martin, Guy P Hunt, Munishikha Kalia, Ahmad Al Khleifat, Simon D Topp, Christopher E Shaw, Ammar Al-Chalabi, Alfredo Iacoangeli

**Author notes:** co-first author (Thomas P Spargo, Sarah Opie-Martin). co-senior author (Ammar Al-Chalabi, Alfredo Iacoangeli).

## Abstract

**Objective:** Variants in the superoxide dismutase (*SOD1*) gene are among the most common genetic causes of amyotrophic lateral sclerosis. Reflecting the wide spectrum of putatively deleterious variants that have been reported to date, it has become clear that *SOD1*-linked ALS presents a highly variable age at symptom onset and disease duration.

**Methods:** Here we describe an open access web-tool for comparative phenotype analysis in ALS: https://sod1-als-browser.rosalind.kcl.ac.uk/. The tool contains a built-in dataset of clinical information from 1,383 people with ALS harbouring a *SOD1* variant resulting in one of 162 unique amino acid sequence alterations, and from a non-*SOD1* comparator ALS cohort of 13,469 individuals. We present two examples of analyses possible with this tool, testing how the ALS phenotype relates to *SOD1* variants which alter amino acid residue hydrophobicity, and distinct variants at the 94^th^ residue of SOD1 which has six variants sampled at the same position.

**Results and conclusions:** The tool provides immediate access to the datasets and enables bespoke analysis of phenotypic trends associated with different gene variants, including the option for users to upload their own datasets for integration with the server data. The tool can be used to study *SOD1*-ALS as well as an analytical framework to study the differences between other user-uploaded ALS groups and our large reference database of *SOD1* and non-*SOD1* ALS. The tool is designed to be useful for clinicians and researchers, including those without programming expertise, and is highly flexible in the analyses that can be conducted.

## Background

Amyotrophic lateral sclerosis (ALS) is a fatal neurodegenerative disease characterised by dysfunction and death of motor neurons leading to progressive muscle weakness and paralysis^1^. Its clinical presentation can vary greatly. For example, although most patients develop the first symptoms between 55 and 65 years of age onset age, the disease can onset across all stages of adulthood. Similarly, time from symptom onset until death is a median of 3 years for ALS but some people die within a year of onset, and 5-10% of people survive for more than 10 years^2-4^.

A plethora of genetic factors can affect the risk of ALS or its phenotype, and mutations in specific genes can lead to distinct clinical outcomes. For example, a hexanucleotide expansion in the *C9orf72* gene is the most common known cause of ALS and carriers of this mutation typically develop ALS earlier and with faster progressing symptoms than sporadic ALS patients^5-8^. Furthermore, different mutations within the same gene can also lead to distinct forms of the disease. For example, over 180 variants in the superoxide dismutase (*SOD1*) gene^9-11^ have been found in ALS patients. As well as affecting ALS risk, some of these variants have distinct effects on clinical features such as age of onset of motor symptoms and disease duration. For example, p.A5V and p.H44R have a marked effect on disease duration while p.G38R is associated with an early onset^10,12-14^. Being able to characterise how genetic variants affect the clinical phenotype is essential for optimal development and design of healthcare, treatments, and trial stratification. However, the multitude of genetic factors involved in ALS and their rarity are great challenges for their individual study. To address these limitations, focussing on *SOD1* given the recent gene therapy trials^15^, we recently collated data from the literature and specialised ALS centres globally on approximately 15,000 people with ALS, over 1,000 of whom harboured a variant in the *SOD1* gene^10^.

In this paper, we describe a web tool (https://sod1-als-browser.rosalind.kcl.ac.uk/) with upload facilities to allow people to perform comparative and bespoke phenotype analyses using data from a database of almost 15,000 people with ALS without need for informatics proficiency. The tool currently allows users to define and select subgroups of patients with or without variants in *SOD1*, to stratify by individual or groups of *SOD1* variants, and to upload data to combine with our database in the analysis. To show the potential of this tool and how to use it, we present two example case studies which leverage the data from our recent publication which is accessible to all users. The first example builds upon research suggesting that variants affecting protein hydrophobicity promote aggregation of mutant SOD1^16^ and tests how alterations in amino acid hydrophobicity affect the ALS phenotype.

The second example focuses specifically on variation at the 94^th^ amino acid residue of SOD1, which is a site containing multiple reported variants, testing how the phenotype differs for each variant sampled.

## Materials and methods

### Dataset

The tool enables access to a dataset of 14,852 people with ALS, 1,383 of whom harbour a potentially deleterious non-synonymous *SOD1* gene variant (N without *SOD1* variant = 13,469). A total of 162 unique amino acid variants (canonical SOD1 sequence IDs: ENSEMBL = ENST00000270142.11, UniProt = P00441) are represented within these data (see Figure 1; Table S1). The dataset is further described within our previous publication^10^ and a summary of the disease characteristics associated with the 49 variants harboured by at least 5 people is provided on the site.

**Figure 1.**
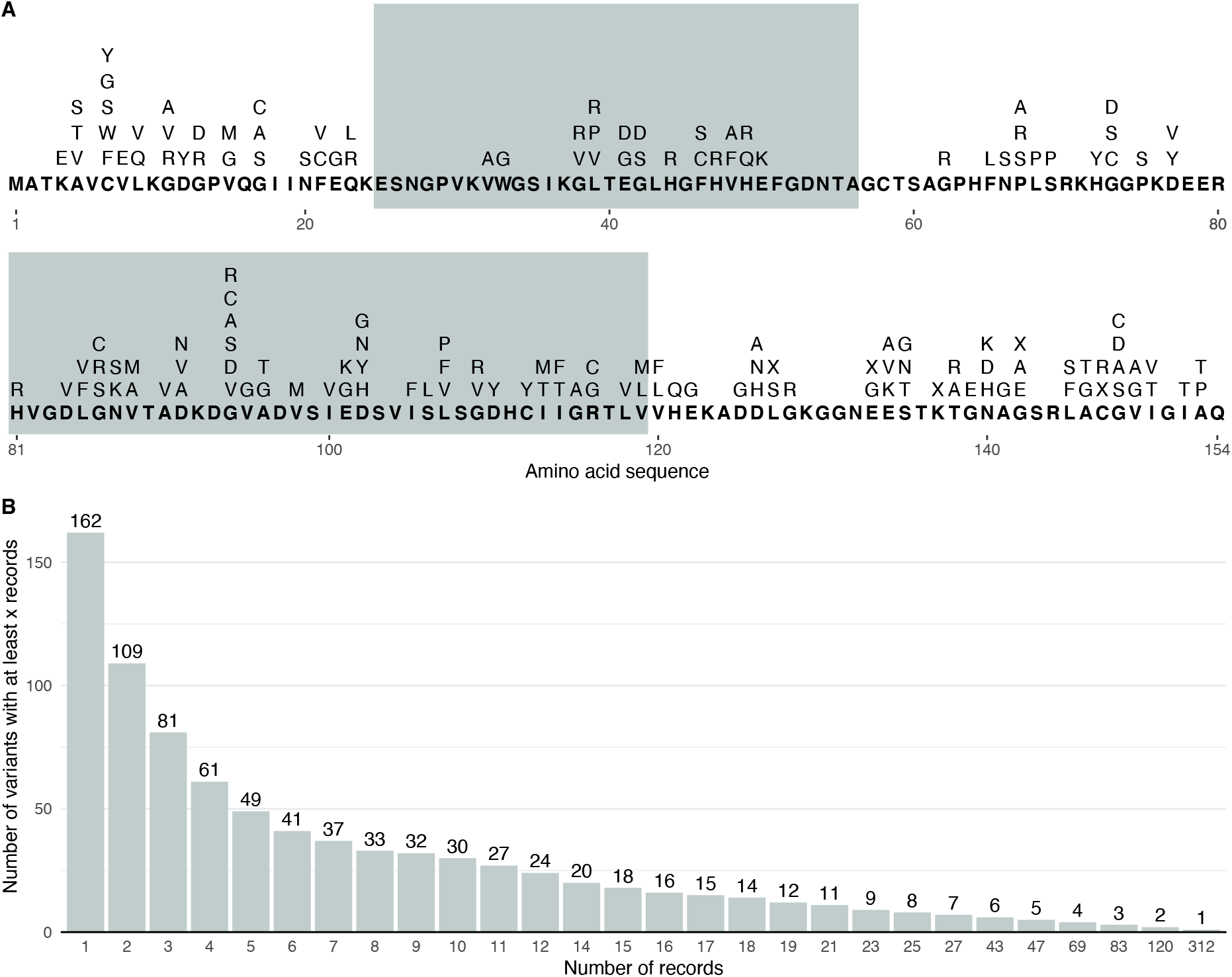
Variant characteristics for the native dataset. **Panel A:** The canonical SOD1 amino acid sequence (bold) and variants recorded at each residue, denoted using IUPAC amino acid nomenclature^30^, where ‘X’ indicates protein truncating variants. Alternating background shading indicates residues encoded from different exons of the SOD1 gene. **Panel B:** The number of variants with at least a certain number of records available across different thresholds.

### Functionality

Survival analysis methods can be performed for (1) age at symptom onset, and (2) disease duration from symptom onset (with a corresponding censor variable indicating survival status). Kaplan-Meier and Cox Proportional-Hazards (CPH) approaches are both implemented and relevant descriptive statistics for the analysed sample are given by strata. Differences between strata in univariate analyses are examined using the log-rank test; global and pairwise log-rank tests are performed when more than two strata are defined.

Analyses using CPH models are performed whenever two or more strata are defined or when a single stratum is specified, and the user selects at least one of several covariates which can be included in the regression model. Available covariates are clinical diagnosis, family disease history, sex, age of onset, site of onset, and sample source (continent of origin). Users can pick which covariates are included in the analysis depending on requirements and associations between selected survival analysis strata and available covariates can be tested.

Various analysis options are provided. The user can model survival for any number of individual SOD1 variants (including a ‘no variant’ option) and variants can be collapsed into groups of interest (including an ‘any other SOD1 variant’ option). We include three pre-defined options for grouping variants: by functional location^10^ in the protein (across the dimer interface, electrostatic loop, zinc loop, and other) or according to the gene exon from which variants are transcribed. The final pre-defined analysis compares people with any SOD1 variant versus the ‘no variant’ group.

Users can further customise the analysis. They can filter by continent of origin and opt to stratify the analysis by sex, family history, site of onset, and country of origin. Time-dependent CPH analyses are also possible, allowing users to define timepoints at which the data are split. This functionality allows time-dependent coefficients to be modelled and enables analysis constrained to a certain timeframe (e.g. only of the first 12 months from symptom onset).

We allow users to upload supplemental data that is appended to the native sample, enriching the analysis possible within the tool. There are no restrictions regarding records that can be uploaded as supplemental data; users may provide data associated with *SOD1* variants both present in and absent from the native data or provide data from other groups of patients, e.g. for variants from other genes. Formatting instructions for supplemental data are provided on the site.

The results of the user’s analyses are presented on the website, and we provide options to (1) download these within an HTML report and (2) download publication-ready versions of the figures produced, with customisable formatting.

### Tool design

The tool is written in the R coding language (R Version 3.6.3) and developed using the R packages (versions) *shiny* (1.6.0), *shinyjs* (2.0.0), *shinycssloaders* (1.0.0), *tidyverse* (1.3.2), *rmarkdown* (2.1.3), *countrycode* (1.3.1), *kableExtra* (1.3.4), *plotly* (4.10.0), and *backports* (1.4.1) ^17-26^. Survival analyses are performed and visualised using *survival* (version 3.3.1), and *survminer* (version 0.4.9)^27,28^.

### Examples of use

Here we present two examples of analyses possible within this tool. We examined differences in age of onset and disease duration between the strata of each example using Kaplan-Meier analyses and the log-rank test, and CPH models with robust variance estimation as implemented by *coxph* were applied to examine differences between strata before and after controlling for possible covariates. In the CPH models, we controlled for sex and age of onset when analysing disease duration, and sex only when analysing age of onset.

Case study 1 examined whether changes to amino acid hydrophobicity influenced age of ALS onset or disease duration from onset until death. Amino acids were grouped into three hydrophobicity categories^29^: hydrophobic (Amino acid IUPAC code^30^: F, M, I, L, V), hydrophilic (D, E, H, K, R, N, Q), and intermediate (Y, W, P, G, A, S, T, C). Variants resulting in an amino acid substitution were then categorised based on the hydrophobicity group of the wild type and mutant amino acid; Table S1 presents the assignment of groups and data availability across variants. To specifically examine the consequence of changes in hydrophobicity, three sets of analyses were conducted, each respective to variants occurring in residues that are hydrophilic, intermediate, or hydrophobic in the wild type protein. In each analysis, variants resulting in altered hydrophobicity were compared relative to variants where the mutant and wild type amino acids remained in the same hydrophobicity group. The p.A5V variant was excluded from these analyses since it is characterised by a particularly aggressive phenotype and accounted for the majority of records (n = 312) in the ‘intermediate to hydrophobic’ category. A broader hydrophobicity analysis across all groups was also conducted.

Case study 2 examined trends associated with variation at the 94^th^ amino acid residue of SOD1, coding for a glycine in the wild type protein. Six variants were present at this locus. We first analysed differences in age of onset and disease duration associated with having any p.G94 variant vs any other SOD1 variant. Second, we compared p.G94 variants individually to non-p.G94 variants, aggregating across p.G94R, p.G94S, and p.G94V since they each contained fewer than 5 records.

Table 1 summarises characteristics of the data from both case studies.

**Table 1.** Data summary for case studies.: SE = standard error; CI = confidence interval.

| Case study | Analysis stratum | Total sample size, N | N with disease duration (N censored) | Restricted mean disease duration (months) [SE] | Median disease duration (months) [95% CI] | N with age of onset | Restricted mean age of onset (years) [SE] | Median age of onset (years) [95% CI] |
| --- | --- | --- | --- | --- | --- | --- | --- | --- |
| 1: amino acid hydrophobicity | hydrophilic | 118 | 106 (36) | 157.93 [22.5] | 85 [66, 125] | 115 | 47.96 [1.08] | 47 [44, 51] |
|  | hydrophilic to intermediate | 227 | 170 (65) | 147.09 [14.28] | 96 [84, 123] | 215 | 49.58 [0.93] | 49 [47, 50.47] |
|  | hydrophilic to hydrophobic | 17 | 15 (4) | 96.59 [29.31] | 65 [39, -] | 17 | 49.77 [2.02] | 48 [46.85, 56] |
|  | hydrophobic | 161 | 127 (51) | 109.18 [17.09] | 74 [50, 109] | 152 | 49.46 [0.92] | 50 [48, 53] |
|  | hydrophobic to intermediate | 221 | 154 (34) | 85.22 [10.29] | 45 [30, 72] | 202 | 50.82 [0.87] | 49.5 [48, 51.19] |
|  | hydrophobic to hydrophilic | 9 | 8 (0) | 26.37 [9.69] | 18 [10, -] | 9 | 50.78 [2.73] | 54 [49, -] |
|  | intermediate | 177 | 135 (36) | 80.11 [14.09] | 38.4 [24, 53] | 173 | 47.04 [1.03] | 46 [44, 49] |
|  | intermediate to hydrophilic | 96 | 76 (15) | 101.59 [15.36] | 45 [32, 84] | 90 | 49.06 [1.49] | 48.5 [45, 51] |
|  | intermediate to hydrophobic | 38 | 29 (9) | 120.85 [30.43] | 84 [56, 168] | 37 | 46.3 [2.26] | 47 [43, 53] |
| 2: p.G94 amino acid residue analysis | non-p.G94 SOD1 variants | 1320 | 1034 (248) | 88.43 [4.83] | 37.59 [30, 44] | 1252 | 49.07 [0.36] | 49 [48, 50] |
|  | Any p.G94 variant | 63 | 52 (10) | 75.21 [17.08] | 32 [26, 53] | 63 | 46.06 [1.81] | 45 [39, 49] |
|  | p.G94A | 27 | 26 (1) | 33.44 [6.55] | 22 [19, 32] | 27 | 48.7 [3.17] | 48 [43, 61] |
|  | p.G94C | 14 | 9 (4) | 221.68 [86.97] | 235.4 [42.84, -] | 14 | 40.46 [2.42] | 37.5 [35, 51] |
|  | p.G94D | 15 | 14 (5) | 55.56 [8.64] | 53 [31, -] | 15 | 49.73 [3.78] | 51 [45, 63] |
|  | p.G94R | 1 | 1 (0) | 55 [0] | 55 [-, -] | 1 | 34 [0] | 34 [-, -] |
|  | p.G94S | 2 | 0 (-) | - | - | 2 | 37.5 [5.3] | 37.5 [30, -] |
|  | p.G94V | 4 | 2 (0) | 114.5 [65.41] | 114.5 [22, NA] | 4 | 41.25 [1.75] | 41 [37, -] |

## Results

### Amino acid hydrophobicity analysis

In case study 1, we examined how the ALS phenotype varied by changes in amino acid hydrophobicity. Across all amino acid substitutions sampled: 42.86% were variants which remained in the same hydrophobicity category as wild type SOD1; 42.11% were variants with a hydrophilic or hydrophobic amino acid in the wild type and an intermediate amino acid in the mutant protein; 12.59% were variants with an intermediate amino acid becoming hydrophilic or hydrophobic; and 2.44% were variants with substitutions from hydrophilic to hydrophobic amino acids or vice versa (see Table 1).

Age at symptom onset appeared roughly comparable across variants in all categories of the hydrophobicity analyses (see Table 2; Table S2; Figure 2), with all groups having a restricted mean age of onset between 46 and 51 years (Table 1).

**Table 2.** Inferential statistics for survival analyses across case studies. Bold values denote nominal p-values < 0.05. ^*^ controlling for sex in the age of onset analysis and for sex and age of onset in the disease duration analysis. ^#^Hazard ratios greater than 1 indicate earlier age of onset/shorter disease duration in the non-reference group. ^†^No p.G94S variants were available for the disease duration analysis. CPH = Cox Proportional-hazards

| Analysis | Case study (reference group) | Analysis stratum | Hazard ratio [95% confidence interval] # |  | P-value of difference between stratum and reference group |  |
| --- | --- | --- | --- | --- | --- | --- |
|  |  |  | Unadjusted | Adjusting for covariates* | Log-rank test | CPH model |
| Age of onset | 1 (hydrophobic) | hydrophobic to intermediate | 0.825 [0.675, 1.01] | 0.822 [0.673, 1] | 0.097 | 0.0546 |
|  |  | hydrophobic to hydrophilic | 1.01 [0.677, 1.52] | 1.01 [0.682, 1.49] | 0.784 | 0.97 |
|  | 1 (intermediate) | intermediate to hydrophobic | 1.09 [0.792, 1.49] | 1.1 [0.798, 1.51] | 0.670 | 0.57 |
|  |  | intermediate to hydrophilic | 0.851 [0.657, 1.1] | 0.851 [0.658, 1.1] | 0.219 | 0.22 |
|  | 1 (hydrophilic) | hydrophilic to intermediate | 0.819 [0.659, 1.02] | 0.819 [0.657, 1.02] | 0.097 | 0.0734 |
|  |  | hydrophilic to hydrophobic | 0.995 [0.702, 1.41] | 0.994 [0.702, 1.41] | 0.979 | 0.974 |
|  | 2 (non-p.G94 SOD1 variants) | Any p.G94 variant | 1.12 [0.831, 1.52] | 1.12 [0.827, 1.52] | 0.344 | 0.466 |
|  |  | p.G94A | 0.857 [0.561, 1.31] | 0.836 [0.544, 1.28] | 0.436 | 0.415 |
|  |  | p.G94C | 2.4 [1.21, 4.76] | 2.48 [1.28, 4.83] | **6.73x10<sup>-4</sup>** | **7.39x10<sup>-3</sup>** |
|  |  | p.G94D | 0.891 [0.57, 1.39] | 0.92 [0.593, 1.43] | 0.659 | 0.708 |
|  |  | p.G94R/S/V | 3.49 [2.27, 5.36] | 3.32 [2.14, 5.15] | **5.66x10<sup>-4</sup>** | **9.34x10<sup>-8</sup>** |
| Disease duration | 1 (hydrophobic) | hydrophobic to intermediate | 1.4 [1.06, 1.85] | 1.32 [1, 1.75] | **0.0202** | 0.0503 |
|  |  | hydrophobic to hydrophilic | 4.36 [2.01, 9.48] | 5.28 [2.87, 9.72] | **1.25x10<sup>-5</sup>** | **9.19x10<sup>-8</sup>** |
|  | 1 (intermediate) | intermediate to hydrophobic | 0.63 [0.41, 0.968] | 0.6 [0.373, 0.966] | 0.0576 | **0.0355** |
|  |  | intermediate to hydrophilic | 0.719 [0.514, 1.01] | 0.686 [0.497, 0.949] | 0.0627 | **0.0228** |
|  | 1 (hydrophilic) | hydrophilic to intermediate | 0.924 [0.676, 1.26] | 0.814 [0.591, 1.12] | 0.619 | 0.209 |
|  |  | hydrophilic to hydrophobic | 1.53 [0.83, 2.8] | 1.4 [0.753, 2.58] | 0.296 | 0.289 |
|  | 2 (non-p.G94 SOD1 variants) | Any p.G94 variant | 1.07 [0.823, 1.4] | 1.14 [0.887, 1.46] | 0.642 | 0.308 |
|  |  | p.G94A | 1.75 [1.35, 2.28] | 1.73 [1.34, 2.25] | **5.95x10<sup>-3</sup>** | **3.00x10<sup>-5</sup>** |
|  |  | p.G94C | 0.446 [0.197, 1.01] | 0.518 [0.236, 1.13] | 0.0672 | 0.100 |
|  |  | p.G94D | 0.89 [0.589, 1.35] | 0.946 [0.65, 1.38] | 0.748 | 0.771 |
|  |  | p.G94R/S/V† | 0.85 [0.415, 1.74] | 0.907 [0.375, 2.19] | 0.788 | 0.828 |

**Figure 2.**
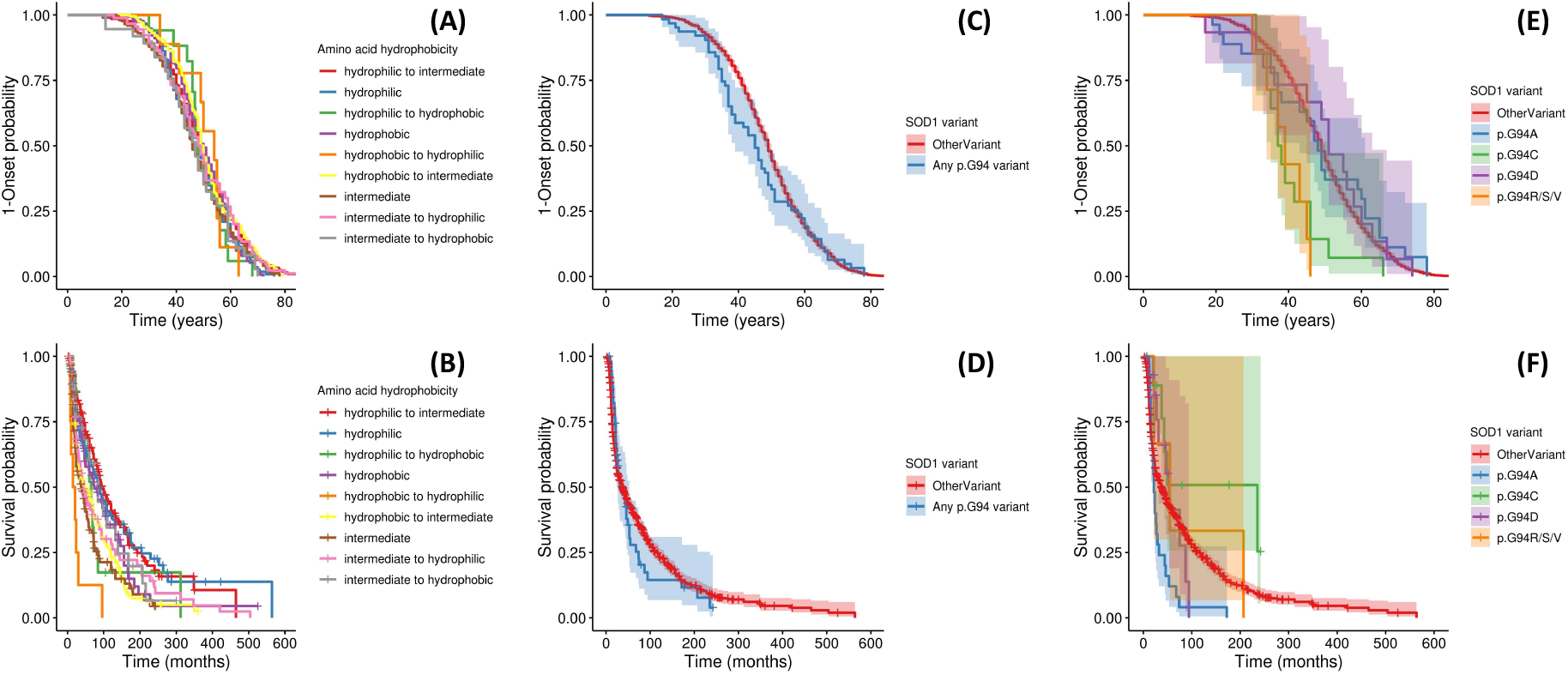
Kaplan-Meier survival curves for age of onset and disease duration analyses. Analysis shown: **Panels A-B:** trends associated with wild type and variant amino acid hydrophobicity; **Panels C-D:** Any SOD1 p.G94 variant versus non-p.G94 SOD1 variants (OtherVariant). **Panels E-F:** individual p.G94 variants versus non-p.G94 SOD1 variants. Panels A, C, and E are for age of onset analysis, and B, D, and F describe disease duration. Panels A and B display all hydrophobicity groups in a single figure for each analysis and confidence intervals are not displayed to maximise visual clarity; Figure S1 visualises trends in age of onset and disease duration for these groups after stratifying across panels according to the hydrophobicity group of the wild type residue.

Disease duration analysis (see Table 2) however, suggested that alterations in amino acid hydrophobicity may affect disease prognosis following onset. Analysis of variants affecting residues which are hydrophobic in wild type SOD1, indicated that disease duration was shorter in substitutions to hydrophilic amino acids (p-value: log-rank test = 1.25×10^−5^; CPH model = 9.19×10^−8^) and that substitution into intermediate amino acids also tended towards shorter disease duration (p-value: log-rank test = 0.0202; CPH model = 0.0503). The median [95% CI] disease duration was 74 [50, 109] months for hydrophobic to hydrophobic substitutions, 45 [30, 72] for hydrophobic to intermediate, and 18 [10, NA] for hydrophobic to hydrophilic.

Among variants occurring in intermediate residues of wild type SOD1, becoming either hydrophilic or hydrophobic was associated with longer disease duration. The median disease duration [95% CI] for mutations of intermediate to intermediate amino acids was 38.4 [24-53] months, under half that of intermediate to hydrophobic (84 [56-168]) and shorter than intermediate to hydrophilic (45 [32, 84]) substitutions.

Analysis of variants in hydrophilic residues of wild type SOD1 did not identify clear differences in disease duration between substitutions which remained hydrophilic and those which became intermediate (p-value: log-rank test = 0.619; CPH model = 0.209) or hydrophobic (p-value: log-rank test = 0.296; CPH model = 0.289). The median disease duration [95% CI] for hydrophilic to hydrophilic substitutions was 85 [66, 125] months, trending towards being shorter than in hydrophobic to intermediate (96 [84, 123]) and longer than in hydrophilic to hydrophobic (65 [39, NA]) substitutions.

Table S3 presents an additional CPH model comparing all hydrophobicity groups relative to substitutions in residues with intermediate to intermediate amino acid substitutions. The analysis indicated that disease duration was shortest in this and the hydrophobic to hydrophilic substitution groups.

### p.G94 amino acid residue analysis

In case study 2, we examined trends associated with variation in the 94^th^ SOD1 residue. p.G94A was the most frequent variant at this locus and 5 other variants occurred in the dataset (see Table 1). This case study showed variant-specific trends in age of onset and disease duration, which were not discernable when aggregating across p.G94 variants, when compared with non-p.G94 SOD1 variants (see Table 2; Figure 2).

Age of onset was earlier than in the non-p.G94 SOD1 variant reference category only in the p.G94C (p-value: log-rank test = 6.73×10^−4^; CPH model = 7.39×10^−4^) and p.G94R/S/V (p-value: log-rank test = 5.66×10^−4^; CPH model = 9.34×10^−8^) groups; this difference appears considerable since the median age of onset for non-p.G94 SOD1 was over 10 years later than median onset in these two groups (see Table 1; Figure 2(E)).

The disease duration analysis indicated that only the p.G94A variant was associated with shorter time to death (p-value: log-rank test = 5.95×10^−3^; CPH model = 3.00×10^−5^). Inspection of hazard ratios suggests that p.G94C trended towards longer disease duration compared to non-p.G94 variants even after controlling for age of onset and sex (p-value: log-rank test = 0.0672; CPH model = 0.100). Although the median disease duration was longer for variants in p.G94D and the p.G94R/S/V variant groups, data were insufficient to test the association.

## Discussion

We have developed a web-tool to facilitate bespoke investigations of the impact of *SOD1* gene variants upon the ALS phenotype, using survival analysis approaches. We have provided two examples of this tool’s utility, examining differences in ALS age at symptom onset and disease duration according to (1) variants of varying impact upon residue hydrophobicity across SOD1 and (2) distinct variants at the 94^th^ SOD1 residue.

This online facility has key benefits for research on the heterogenous ALS phenotype. First, it permits a user-friendly interface for performing survival analysis, with various options for customisation in accordance with the user’s needs. Second, it provides access to a large in-built *SOD1*-ALS cohort and non-*SOD1* comparator population, which can be further enriched if users provide their own supplementary data.

The hydrophobicity analysis suggested that substitution variants altering residue hydrophobicity from hydrophobic to intermediate or hydrophilic, are associated with a shorter disease prognosis compared to variants in residues remaining hydrophobic across wild type and mutant SOD1. This aligns well with evidence that altered hydrophobicity promotes aggregation of the SOD1 protein^16^, and may reflect greater destabilisation and misfolding of SOD1 when variants cause more extreme alterations in hydrophobicity^31-33^. Interestingly, variants of intermediate to intermediate amino acid substitutions were characterised by particularly short disease duration.

Hydrophobic to hydrophilic amino acid substitutions and vice versa were, notably, infrequent relative to other substitutions. Given that these would represent the most extreme hydrophobicity alterations this could indicate a potential survivorship bias and that these substitutions may be sufficiently deleterious to be evolutionarily suppressed. This appears reasonable since SOD1 is highly conserved, with deficiency being linked to severe and early onset phenotypes^34-36^, and on the basis of variants in these hydrophobicity groups being entirely absent from the gnomAD v2.1.1 population database^37^ (see Table S4).

Analysis of the variants at p.G94 emphasised the extent to which individual SOD1 variants differentially influence the phenotype. Grouping together all p.G94 variants suggested age of ALS onset and disease duration is comparable across people with variants that variation at this residue and those with non-p.G94 SOD1 variants. Only by examining variants individually did we observe that p.G94A was associated with shorter, and p.G94C trended towards longer, disease duration than non-p.G94 *SOD1*-ALS. Likewise, p.G94C and the aggregation of p.G94R, p.G94S, and p.G94V were indicative of substantially earlier age of onset. These findings are consistent with the results of our previous analysis of *SOD1*-ALS, emphasising distinction between trends in age of onset and disease duration across individual variants^10^. They highlight particularly the importance of making available resources to allow variant-level analyses of the ALS phenotype associated with variation in *SOD1*.

The tool is not without limitation. Most notable is that a number of the 162 SOD1 variants sampled are harboured by very few individuals and thus are not sufficient for individual variant analysis with the native dataset alone. However, this issue can be somewhat circumvented by aggregating rarer variants into a single analysis stratum, and by the possibility of increasing the dataset with user-supplied data.

Certain considerations apply when providing supplementary data to the tool. Firstly, CPH models may only include covariates that are available in the native dataset. Second, records from supplementary data may overlap with native dataset. To reduce this possibility, the tool will automatically flag any people among the supplementary dataset who may be a duplicate of a person in the native data, checking for matches by country of origin, SOD1 amino acid change (if the user indicates that one is present), age of onset, site of onset, sex, and disease duration (if not censored). Users can also consult the cohort description provided and contact ALSoD (https://alsod.ac.uk/; ^38^) with any concerns.

Overall, the open-access web-utility we provide (https://sod1-als-browser.rosalind.kcl.ac.uk/) has a potentially substantial benefit for ALS disease research and direct translational use for the design of patient stratification approaches, as well as being useful for mutation adjudication committees needing to make decisions on likely disease course with limited data. It permits an array of analysis options which can be readily implemented by users without any programming knowledge, and can be enriched by the provision of a supplementary dataset. Accordingly, this tool allows clinicians and researchers to circumvent many possible barriers they may otherwise face, for instance, regarding insufficient data availability or in preparing these data for analysis. The potential translational benefit of this tool is substantial, facilitating growth in understanding of the ALS phenotype which may aid the design and implementation of effective healthcare, treatments, and clinical trials.

## Data Availability

All data produced in the present study are available upon reasonable request to the authors

## Data availability

All data associated with this manuscript can be accessed from the web-utility: https://sod1-als-browser.rosalind.kcl.ac.uk/.

## Acknowledgments

Samples used in this research were in part obtained from the UK National DNA Bank for MND Research, funded by the MND Association and the Wellcome Trust. We thank people with MND and their families for their participation in this project. We acknowledge sample management undertaken by Biobanking Solutions funded by the Medical Research Council at the Centre for Integrated Genomic Medical Research, University of Manchester. The authors acknowledge use of the research computing facility at King’s College London, *Rosalind* (https://rosalind.kcl.ac.uk), which is delivered in partnership with the National Institute for Health Research (NIHR) Biomedical Research Centres at South London and Maudsley and Guy’s and St. Thomas’ NHS Foundation Trusts, and part-funded by capital equipment grants from the Maudsley Charity (award 980) and Guy’s and St. Thomas’ Charity (TR130505). The authors also acknowledge the use of the CREATE research computing facility at King’s College London^39^. We also acknowledge Health Data Research UK, which is funded by the UK Medical Research Council, Engineering and Physical Sciences Research Council, Economic and Social Research Council, Department of Health and Social Care (United Kingdom), Chief Scientist Office of the Scottish Government Health and Social Care Directorates, Health and Social Care Research and Development Division (Welsh Government), Public Health Agency (Northern Ireland), British Heart Foundation and Wellcome Trust.

## Supplemental online material

Supplementary Tables S1-S4 are available, and captioned, in the file: SOD1browser_supplementary_tables.xlsx

Figure S1. Kaplan-Meier survival curves for age of onset and disease duration analyses of trends associated with wild type and variant amino acid hydrophobicity. The panels stratify according to the wild type SOD1 hydrophobicity group for the residue containing the variant: **panels A-B** show analysis in hydrophobic residues, **panels C-D** for intermediate residues, and **panels E-F** for hydrophilic residues.

