## Supplementary figures and images for "SOD1-ALS-Browser: a web-utility for investigating the clinical phenotype in *SOD1* amyotrophic lateral sclerosis"

### Supplementary figure

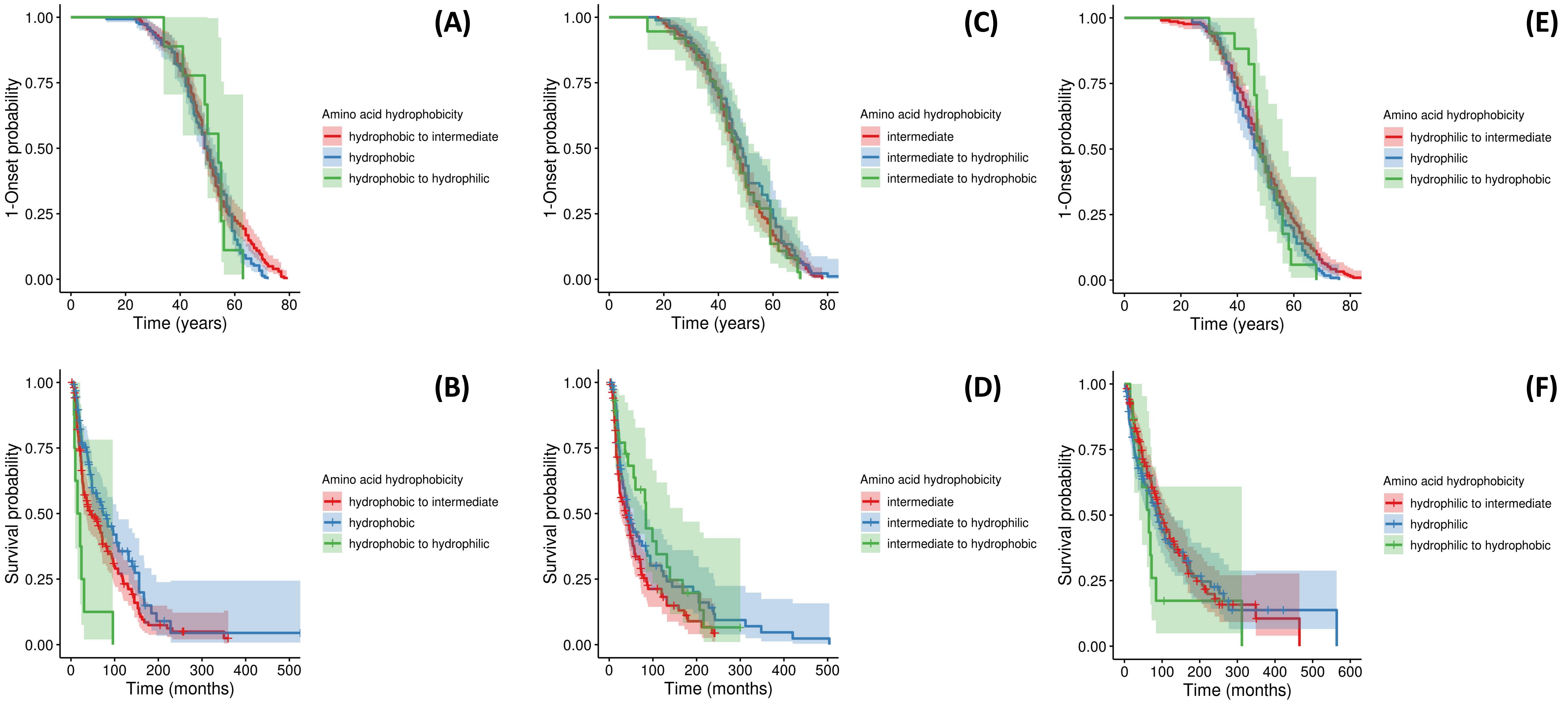
